# Vent-Lock: A 3D Printed Ventilator Multiplexer to Enhance the Capacity of Treating Patients with COVID-19

**DOI:** 10.1101/2020.09.16.20195230

**Authors:** Helen Xun, Christopher Shallal, Justin Unger, Runhan Tao, Alberto Torres, Michael Vladimirov, Jenna Frye, Mohit Singhala, Brockett Horne, Pooja Yesantharao, Bo Soo Kim, Broc Burke, Michael Montana, Michael Talcott, Bradford Winters, Margaret Frisella, Bradley Kushner, Justin M. Sacks, James K. Guest, Sung Hoon Kang, Julie Caffrey

**Affiliations:** Johns Hopkins School of Medicine, Baltimore, MD, 21231; Department of Biomedical Engineering, Johns Hopkins University, Baltimore, MD, 21231; Department of Civil and Systems Engineering, Johns Hopkins University, Baltimore, MD, 21218; Maryland Institute College of Art, Baltimore, MD, 21217; Department of Mechanical Engineering, Johns Hopkins University, Baltimore, MD, 21218; Washington University in St. Louis School of Medicine, St. Louis, Missouri, 63130

## Abstract

Mechanical ventilators are essential to patients who become critically ill from acute respiratory distress syndrome (ARDS), and shortages have been reported due to the novel severe acute respiratory syndrome coronavirus 2 (SARS-CoV-2). We utilized cost-effective, on-demand 3D printing (3DP) technology to produce critical components for a novel ventilator multiplexer system, Vent-Lock, to split one ventilator or anesthesia gas machine between two patients. FloRest, a novel 3DP flow restrictor, provides clinicians control of tidal volumes and positive end expiratory pressure (PEEP), using the 3DP manometer adaptor to monitor pressures. We tested the ventilator splitter circuit in simulation centers between artificial lungs and used an anesthesia gas machine to successfully ventilate two swines. As one of the first studies to demonstrate splitting one anesthesia gas machine between two swines, we present proof-of-concept of a *de novo*, closed, multiplexing system, with flow restriction for individualized patient therapy. Our studies underscore that while possible, ventilator multiplexing is a complicated synergy between machine settings, circuit modification, and patient monitoring. Consequently, ventilator multiplexing is reserved only as a last emergency resource, by trained clinicians and respiratory therapists with ventilator operative experience.

## INTRODUCTION

The novel severe acute respiratory syndrome coronavirus 2 (SARS-CoV-2) has led to a global pandemic resulting in rapid depletion of resources necessary to care for critically ill patients, such as mechanical ventilators and their associated parts. Mechanical ventilators are critical for the treatment of approximately 5-10% of patients with coronavirus disease (COVID-19) who become critically ill from acute respiratory distress syndrome (ARDS)^1^. In the face of the COVID-19 pandemic, it is estimated that there is a global ventilator shortage of 880,000 ^2^. This shortage may disproportionately affect developing countries who suffer from lack of medical infrastructure and resources ^3^, historically resulting in higher mortality rates in pandemics such as the Spanish Flu ^4^. For example, the continent of Africa has limited ventilator capacity, with only 2,000 ventilators across 41 countries ^5^. This capacity deficit is further worsened by the increased need due to the COVID-19 pandemic.

Ventilator shortages occur in resource-rich countries as well. Previous disasters and current projections suggest that hospitals may be operating at 120-160% capacity in the face of a pandemic or national disaster ^6^. Projections suggest that if 20% of the United States population is infected with the virus, there will be significant deficiencies in intensive care unit beds and mechanical ventilators ^6,7^. Furthermore, given the potential for a second wave of infection, epidemiologists predict that if countries continue to lift restrictions used to slow the spread of the virus too early, then a second global peak may result in a further shortage of medical supplies ^8^, ventilators and ventilator associated parts. The Society of Critical Care Medicine shared that clinicians continue to report ventilator shortages in summer of 2020, including 53% of 587 surveyed ICU clinicians did not have enough ventilators and had to use non-standard ventilators or non-invasive devices, and a small percentage declined care due to shortage of ventilators, or placed two patients on one ventilator^9^. Consequently, there is a critical need to address urgent ventilator shortages in the face of the COVID-19 pandemic.

While the introduction of more ventilators, either *de novo* or commercial, could solve the urgent medical needs arising from the COVID-19 pandemic, this solution represents a theoretical ideal that cannot be achieved given the current monetary, time, hospital infrastructure, limited scaling and production capacity, and supply chain constraints ^10,11^, further exacerbated by lack of standardization of parts across brands ^12^. An alternative strategy to quickly increase ventilator capacity as an immediate step in urgent settings is to “split” or multiplex ventilators and anesthesia gas machines. Ventilator multiplexing allows the usage of one machine to ventilate multiple patients and effectively increases the clinical capacity to support urgent needs.

The concept of using one ventilator to support multiple patients during a disaster surge was first published in 2006 by Neyman *et al*.^13^, who reported that four patients could be supported for 12 hours using ventilator equipment and tubing. However, these *in vitro* studies were restricted to patients with similar body habitus and lung compliances. Despite these study limitations, it introduced ventilator multiplexing as a potential solution for emergency situations. In 2017, this proof-of-concept was demonstrated in actual patients during the 2017 Las Vegas shooting ^14^ when a physician supported multiple patients on a single ventilator as a temporary emergency situation until more resources became available. The ability to multiplex ventilators is valuable for global preparedness mechanisms to promptly increase ventilator capacity as an immediate response for disasters, such as trauma surges, natural disasters, or in military frontlines.

Emergency use of ventilator multiplexing is dependent on the dynamic lung states of the patients, including associated lung compliances and airway resistances that drive airflow balance. In the evolving pathologic state of COVID-19 patients with ARDS, an interdependent ventilation system poses many safety concerns. The Society of Critical Care Medicine and other societies in respiratory care issued a joint statement ^15^ summarizing main concerns with ventilator multiplexing ^16^, including the inability to independently monitor and control ventilation parameters (volumes, pressures, rates) critical for ARDS treatment, thus risking adverse outcomes such as underventilation, or ventilator induced lung injury (VILI) such as barotrauma. Additional concerns include ventilator alarm management, disrupted balance of ventilation if a patient has spontaneous breathing, sudden deterioration, kink in the tubing, and viral contamination if breathing circuits between patients are mixed, or the circuit becomes open.

While these barriers exist, they are not insurmountable for emergency ventilator multiplexing. For example, recent engineering solutions have emerged using off-the-shelf medical components that mitigate concerns in ventilator multiplexing by volume or pressure control, and monitoring ^17^. It is optimistic that these circuits present potential solutions for ventilator shortages in emergency situations; however, deployment can be limited due to unstable supply chains that make these off-the-shelf medical components difficult to acquire ^18^, even in developed countries up to six months from inciting events ^19^. Consequently, rapid production of *de novo* ventilator multiplexing solutions are investigated to further address these barriers.

Among multiple modalities for *de novo* manufacturing to address medical equipment shortages, 3D printing has come to the forefront during the COVID-19 pandemic to address critical shortages ^12,20^. 3D printing is a type of additive manufacturing that has emerged in the past decades as a cost-effective, rapid on-demand production modality with broad applications due to its ability to produce intricate and complex geometries from computer-aided designs without tooling and expensive machines ^21^. 3D printing enables faster design and manufacturing processes ^22^, so that it can be utilized in emergency situations to fill gaps in the supply chain ^23^. Specifically, in reference to COVID-19, there have been multiple shortages in ventilator associated equipment and valves, leading healthcare providers to look into different avenues of manufacturing to address the gap in the supply ^20^. Ventilator splitter products which primarily use commercial medical equipment may have limited or unreliable supply ^17^ in urgent situations. We address urgent medical needs arising from unstable supply chains by using 3D printing to rapidly and cost-effectively prototype and test components of our ventilator splitter circuit using biocompatible and sterilizable materials.

In this study, we present Vent-Lock, a *de novo*, ventilator multiplexing system that addresses major concerns with ventilator splitting, and is rapidly produced via 3D printing, thus tapping into a broad international production infrastructure largely unaffected by the pandemic. The Vent-Lock breathing circuit provides clinicians with a way to control, manage and monitor patients split on one ventilator; circuit components allow for the change in individual tidal volumes and positive end expiratory pressure (PEEP), pressure monitoring, and minimized back flow and risk of contamination. Our novel, air-tight, 3DP flow restrictor (FloRest) is designed to provide clinicians with precise control of tidal volumes. We validate the use of Vent-Lock FloRest for both ventilators and anesthesia gas machines, successfully ventilating simulated patients with mismatched lung compliances. Furthermore, we ventilated two swine safely with Vent-Lock, demonstrating the device’s ability to both safely multiplex patients and to evolve anesthesia gas machines (which are more commonly available in developing countries as compared to ventilators) with increased ventilation settings. We share differences in multiplexing of anesthesia gas machines and ventilators, and the impact of ventilator control mode (volume control versus pressure control) on multiplexing, and additional challenges. The novel, *de novo*, 3DP Vent-Lock circuit and FloRest is well positioned to rapidly increase capacity of mechanical ventilators to provisionally meet ventilator shortages due to the COVID-19 pandemic and future pandemics and disasters.

## RESULTS

### Vent-Lock 1+n(1) circuit and components

We validated a 1+n(1) system which can split one ventilator between one standard patient and one or potentially more variable patients (Fig.1). The standard patient ideally has the lowest lung compliance and has minimal components in the circuit to establish low resistance allowing the ventilator to maintain standard function. The standard patient will be ventilated at pressure settings unaltered from that delivered by the ventilator. Additional patients (n) added to the circuit are considered variable patients and can have their tidal volumes and PEEP altered by circuit components. This paper demonstrates use of a ventilator splitter adjusting for 1 control and n=1 variable patients. The 1+n(1) split contains Vent-Lock 3DP parts and commercial parts (Fig. 1). We 3D printed the splitters and the flow restrictors (needle valves). The other parts including the one-way check valves, the filters and the PEEP valves were all commercial parts (fig. S1). The Vent-Lock circuit was designed to be closed circuit and leak-free to minimize risk of aerosolizing viral particles into the surrounding environment.

**Fig. 1.**
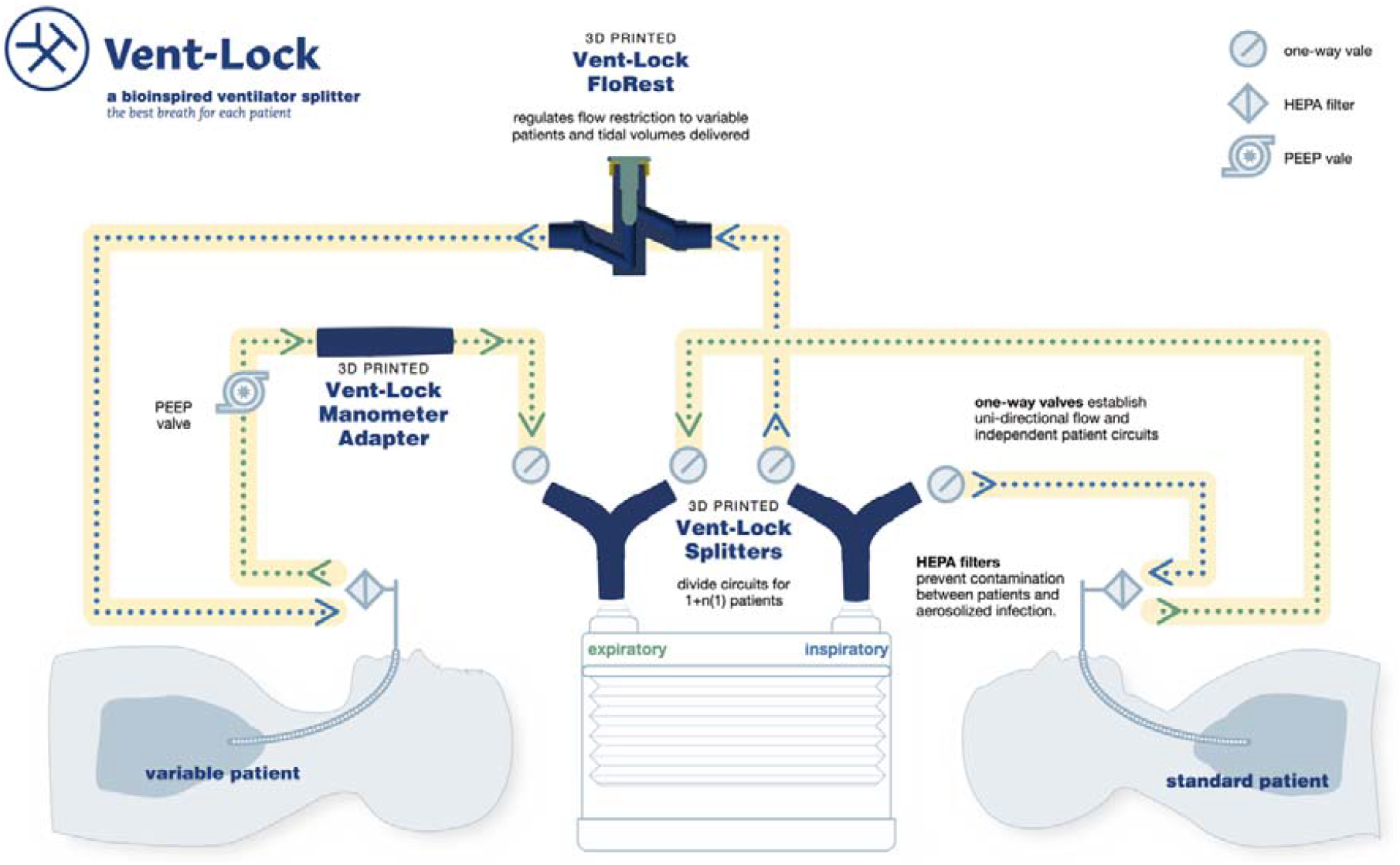
Vent-Lock ventilator multiplexing 1+n(1) circuit and 3DP components. Our 1+n(1) circuit proposes having a standard patient with minimal features, thus are ventilated per ventilator settings. Additional patients added to the circuit will be considered n(1), and will have variable flow and PEEP as controlled by circuit components. Please note that all patients, regardless of standard or variable, have one-way (check) valves and filters.

### Vent-Lock 3DP Flow Restrictor (FloRest)

The Vent-Lock 3DP flow restrictor (Vent-Lock FloRest) is a flow restrictor based on a needle control valve design optimized for low flow rates to offer clinicians robust control over a range appropriate for human ventilation. Vent-Lock FloRest was designed to address the following concerns regarding ventilator “splitting” (15): 1. Volumes would distribute unevenly between patients, 2. PEEP would be difficult to manage per patient, 3. Tidal volumes would be difficult to manage per patient and 4. Adjustment or discontinuation of ventilation to one patient would alter breathing dynamics to other patients.

The goal of FloRest (Fig. 2A) was to allow the clinician to modify the flow rate of air being delivered to the patient, thus providing ranges of clinical tidal volumes and PEEPs with control sensitivity and a reliable linear relationship between closure and flow rate, tidal volume,

**Figure 2.**
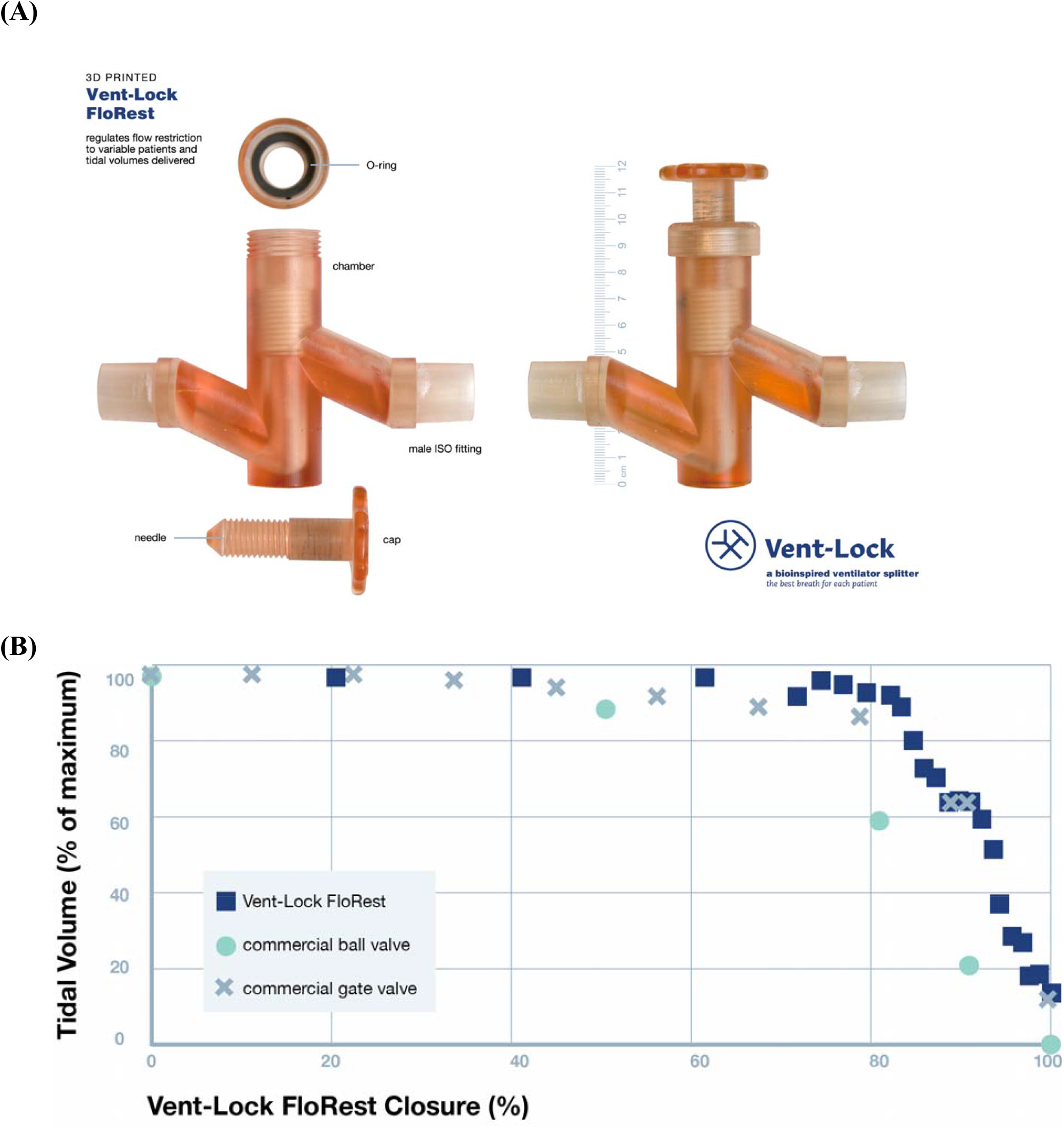
Design and performance of 3DP Flow Restrictor (FloRest). (A) Vent-Lock 3DP Flow Restrictor (FloRest) contains three components. The O-ring between the cap and full-height threaded needle interface at the top of the chamber to maintain air-tight seals. Both ends of Vent-Lock FloRest are male ISO fittings to ensure connection to ventilator tubing. (B) Testing with a ventilator on pressure control using simulation lungs at varying compliances demonstrates that Vent-Lock FloRest provides more control options than commercial ball valves and gate valves, characterized by more available data points corresponding with different tidal volumes delivered.

and pressures. The design emphasizes the minimization of build time and volume by reducing support material use and complex structures for consistent and higher quality printing. These considerations allowed for an air-tight and leak-proof design (fig. S2) and utilization of biocompatible materials that can withstand extended exposure to warm humidified air and sterilizing autoclave environment. Using a particle counter, post and pre-autoclave tests demonstrate significant micron particle reduction after autoclaving (fig. S3).

The needle valve utilizes change in flow momentum, flow path geometry and orifice flow design concepts allowing easy control of flow rate vs pressure drop ratio (i.e. flow coefficient) compared to gate and ball valve concepts (more binary valve concepts). It operates with less total pressure drop over the flow control ranges than typical globe valve concepts. The threading allows for control over the flow rates and offers the clinician the ability to make fine adjustments to the flow within the range of control. We used a gasket-inspired design featuring an O-ring (E1000-212/AS568-212, O-Rings EPDM, FDA EPDM, Marco Rubber &Plastics, Seabrook, New Hampshire, USA) seated between the needle and chamber to ensure airtightness, thus reducing the risk of aerosolizing the virus into the surrounding environment. Final features of FloRest (Fig. 2A) included sealing to the external environment using unthreaded upper needle shafts for smoother interfaces between cap O-ring and needle during operation of valve and a delayed start in needle threading to provide earlier range of flow control and to allow for safe operation of valve by preventing full occlusion of flow to patient by clinician.

The FloRest has advantages compared with commercial valves in terms of the controllability, biocompatibility, and sterilizability. The FloRest had similar range of control compared to commercial gate valves (#P20034 PVC SCH 40 ½-in FNPT Ball Valve; G300 Lead Free Brass Gate Valve, American Valve, Greensboro, North Carolina, USA) (Fig. 2B). However, the Vent-Lock FloRest provides more control options than commercial valves, characterized by more points available for the clinician to choose from corresponding to different tidal volumes delivered. Furthermore, FloRest is produced with biocompatible, nontoxic materials that can be safely sterilized, as compared to commercial ball valves with untested biocompatibility and unknown sterilization protocols. Vent-Lock FloRest can be produced at an estimated $3.50 per device in 3-hour 40 min print and process time via fused deposition modeling (FDM) (e3d, BigBox3D Ltd, Oxfordshire, UK; Little Monster, Tevo 3D Electronic Technology Co. Ltd, Zhanjiang, China) using PETG (PETG 3D Printer Filament, FilaMatrix, Virginia, USA). With stereolithography (SLA; Form 2, Form 3, or Form 3B, Formlabs), it costs approximately $25, and 16 hours production time with a 50 micron build layer height resolution, using surgical guide resin (Surgical Guide, Formlabs). We demonstrate that the FloRest is leak proof through air volume testing (fig. S2).

### Vent-Lock 3DP Flow Restrictor (FloRest) control of tidal volumes and PEEP

We tested the use of Vent-Lock FloRest in the Johns Hopkins Medicine Simulation Center (JHMSC) to confirm the following: 1) Allowing volumes to be distributed evenly between patients, 2) variable patient control of PEEP, 3) variable patient tidal volume control and 4) changes in the variable patient breathing settings does not alter breathing dynamics to the standard patient.

We tested the Vent-Lock multiplexing system using a 1+1 split patient circuit (Fig. 1). We used one ventilator (Puritan Bennett 840 Ventilator System, Avante Health Solutions) to ventilate two patients with different lung compliances of 20 cmH_2_O and 50 cmH_2_O. We first tested using a pressure control mode, with inspiratory pressures set at 25 cmH_2_O (additional ventilator settings available in Table S2). The Vent-Lock FloRest allowed adjustment of tidal volumes delivered to patients between 7-turns to 9-turns (fully closed), corresponding to 21.5% of tidal volume control range and 2-⅛ turning range control (Fig. 3A). It allowed control of 85.7% of the total range of delivered tidal volumes (compared to initial variable patient tidal volume) with negligible change in tidal volume delivery to the standard patient (range: 99.86% and 103.2% initial standard patient tidal volume, mean: 102.1% ± 0.98%). We note that the total expiratory volume reported by the ventilator trends with tidal volume delivered to the variable patient (Fig. 3A) and the peak inspiratory pressures (PIP) of the variable patient and ventilator peak inspiratory volumes also correspondingly decrease with decreases in tidal volume (Fig. 3B,C), while peak inspiratory volumes remain stable for the standard patient and peak end expiratory volumes remain stable for both patients during these changes.

**Fig. 3.**
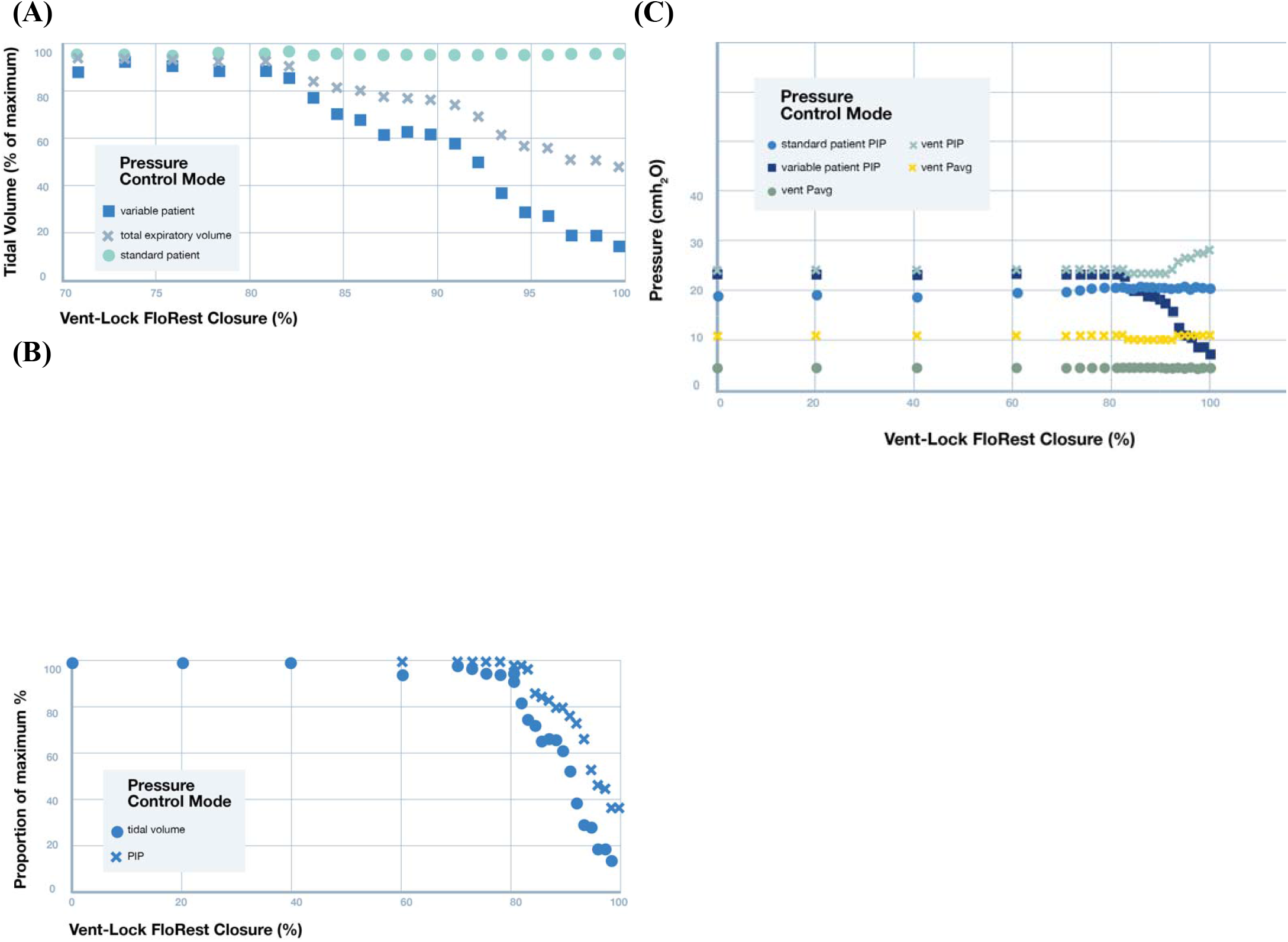
Pressure control mode: testing of Vent-Lock ventilator multiplexor with a ventilator on pressure control mode ventilating two simulated patients with different lung compliances. (A) When used with a ventilator on pressure control, Vent-Lock FloRest is capable of controlling tidal volumes delivered to the variable patient per turn, with no chang to tidal volumes delivered to the standard patient. (B) PIP of the standard patient remains stable, despite changin PIP of the variable patient. Using a ventilator on pressure control, we determine the changes in standard and variable patient breathing pressures with closure of the Vent-Lock FloRest, and the ventilator reported pressures. The positive inspiratory pressure (PIP) of the variable patient decreases with closure, while the standard patient PIP, an ventilator reported average breathing pressures (P_avg_), and PEEP remain constant. We do note an increase in the ventilator reported PIP. (C) With the ventilator on pressure control, changes to tidal volume delivered to patients using Vent-Lock FloRest demonstrates a corresponding change in peak inspiratory pressures (PIP).

We repeated Vent-Lock 1+1 multiplexing patient circuit with the ventilator on volume control mode to deliver a total of 2L of volume, corresponding to approximately 600 mL of tidal volume per patient (additional ventilator settings available in table S1). We note that turning of FloRest on the variable patient resulted in decrease of both tidal volumes and PIP (Fig. 4A,B). However, this was accompanied with an increase in tidal volume delivery and PIP to the standard patient (Fig. 4A,B), with relatively stable ventilator reported average pressures (Vent P_avg_) and PEEP (Fig. 4B). Thus, unlike in pressure control mode where control of delivery to the variable patient was independent of the standard patient, flow restriction in the volume control mode resulted in the modification of the ratio of tidal volumes delivered (Fig. 4C, standard/variable patient tidal volume ratios).

**Figure 4.**
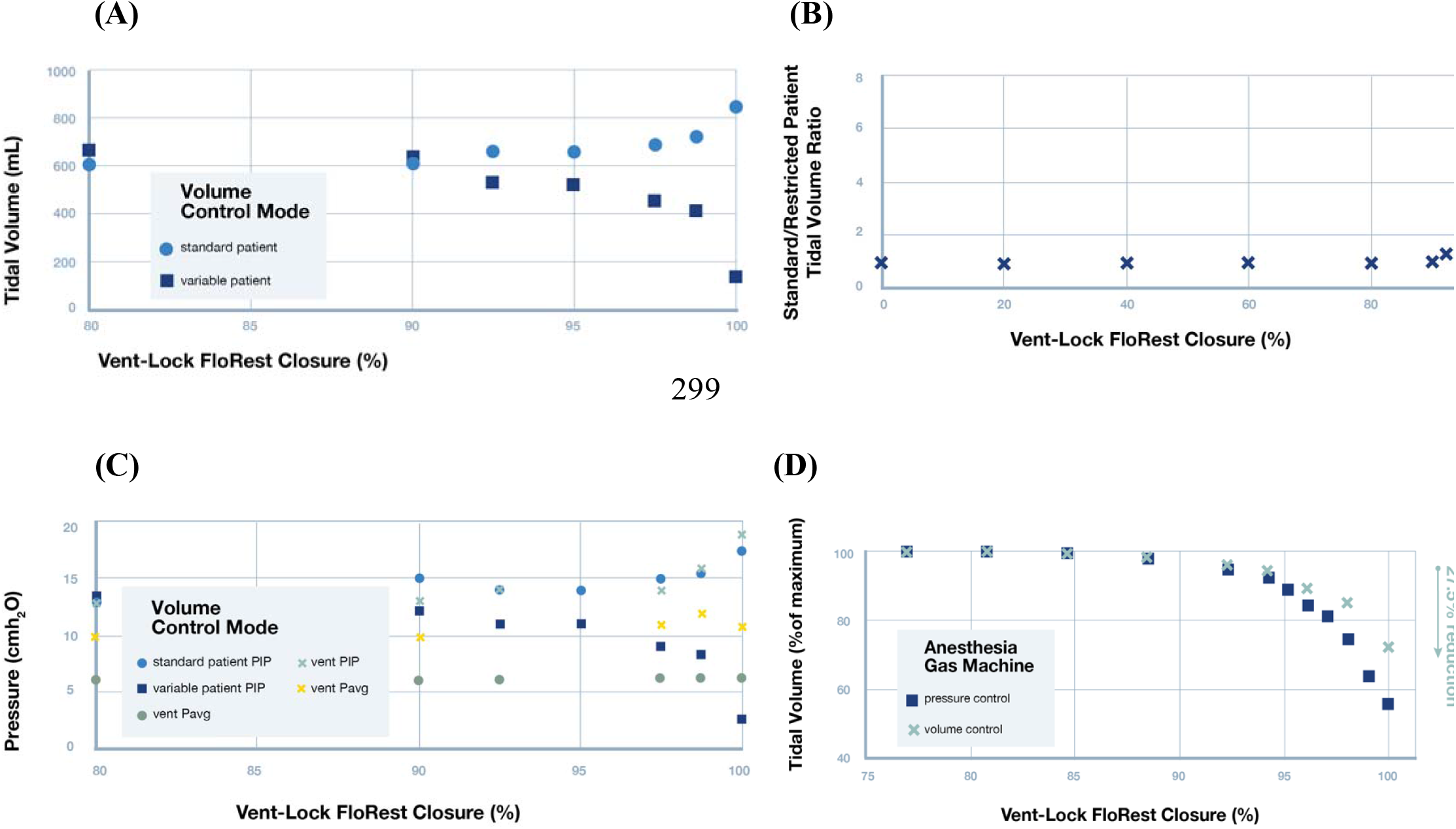
Volume control mode: testing of Vent-Lock ventilator multiplexer with a ventilator or anesthesia gas machine on volume control mode ventilating two simulated patients with different lung compliances. (A) With the ventilator on volume control, decreases in variable patient tidal volumes result in increases in tidal volumes delivered to the standard patient. This indicates that in volume control mode, patient ventilation circuits are interdependent, and changes in one patient effects the other. (B) The ratios of the patient’s tidal volumes (standard patient/variable patient) per closure of the Vent-Lock FloRest with ventilators on volume control. (C) On ventilator volume control and with Vent-Lock FloRest closure, the changes in peak inspiratory pressure (PIP) of the standard patient and variable patient reflect that of tidal volume changes, while the ventilator reported PIP and P_avg_ increase, and PEEP remains stable. (D)Vent-Lock FloRest was tested at Washington University in St. Louis using anesthesia gas machines. The FloRest can be used to control delivered tidal volumes to the variable patient on both pressure and volume control on anesthesia gas machines.

We replicated results using anesthesia gas machines (North American Drager Narkomed 2a, Ardus Medical; GE Aestiva 5 7900, Datex Ohmeda) at an alternate test site (Washington University in St. Louis, St. Louis, Missouri, USA). The 1+1 circuit was tested with the Vent-Lock FloRest on the variable patient. On both pressure control and volume control settings, the Vent-Lock FloRest demonstrated control of tidal volume delivered to the variable patient with stable tidal volumes delivered to the standard patient. Pressure control allowed slightly greater range of control (Fig. 4D, reduction of 43.9% tidal volume at close, compared to 27.5% reduction of tidal volume at close with volume control)

### Real-time pressure reporting with Vent-Lock manometer adaptors

To facilitate continuous monitoring of pressures we designed a manometer adaptor that allows clinicians to either spot-check pressures or continuously monitor with the use of standard, disposable manometers such as those found on bag-valve-masks. The manometer adaptor can be added in the circuit at any point and is designed to accurately reflect breathing pressures, such as PIP and PEEP. We demonstrate that the manometer accurately reflects real-time pressures when incorporated in the circuit (Fig. 5A). In a 1+1 circuit with the ventilator on pressure control, the PEEP setting was incrementally increased, and associated ventilator detected PEEPs and Vent-Lock manometer reported pressures were recorded. The Vent-Lock manometer reported pressures were equivalent to the PEEPs (Figure 5B). We also conducted blind tests, where one researcher set the PEEP on the ventilator and a second researcher (blinded to ventilator PEEP settings) reported PEEP as reported by the Vent-Lock manometer. The second researcher consistently and accurately reported all test values between 0 cmH_2_O to 50 cmH_2_O, in 5 cmH_2_O increments, with total ten trials with no error.

**Fig. 5.**
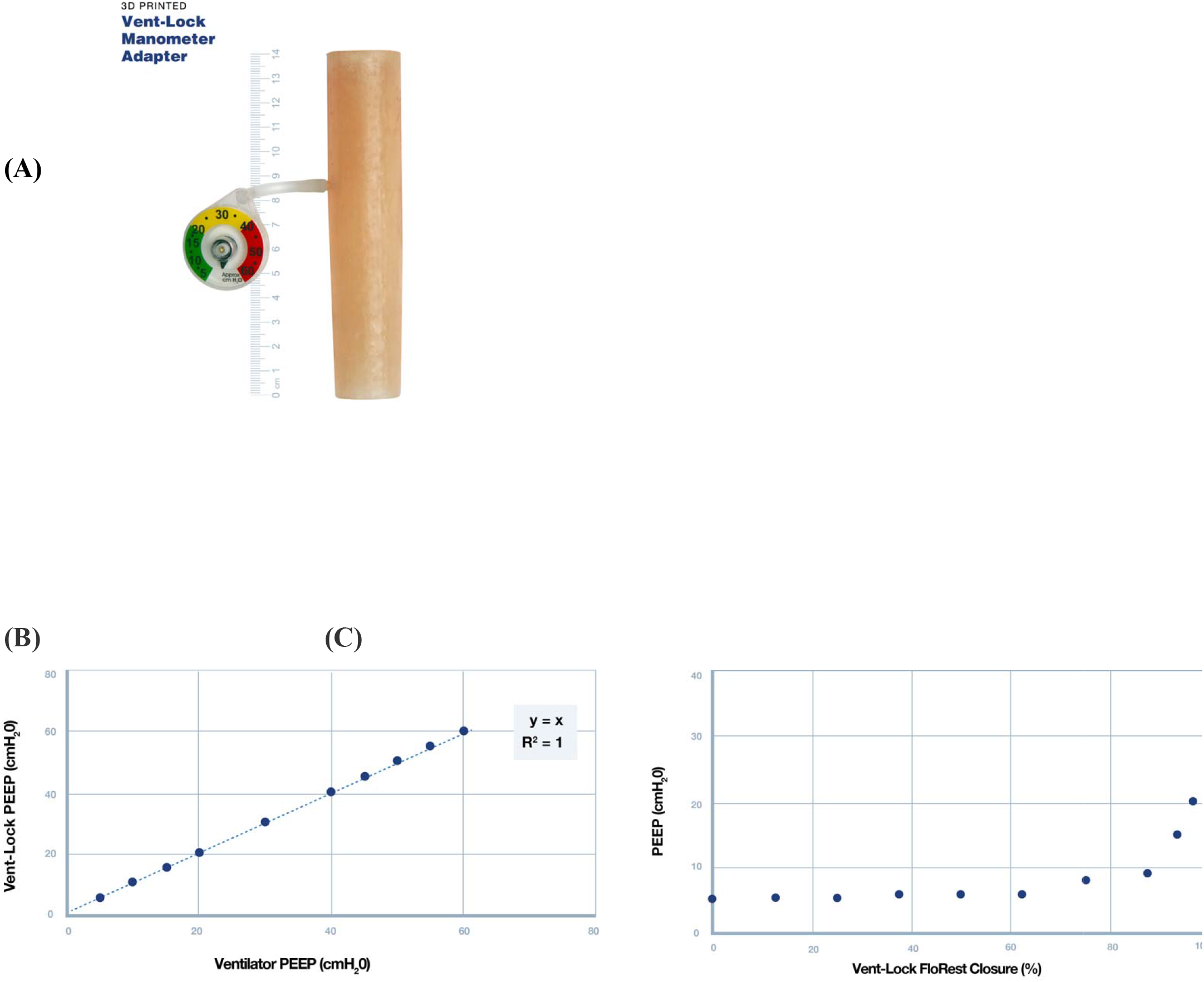
Vent-Lock manometer adaptor. (A) The Vent-Lock manometer adaptor is incorporated in the split circuit, and allows for the attachment of disposable manometers, thus provides accurate, real time readings of pressures. (B) When placed on the expiratory limb, the Vent-Lock manometer adaptor accurately reflects PEEP as set by th ventilator and as reported by the ventilator. (C) We use the Vent-Lock manometer to report the PEEP of the variable patient, as adjusted by the Vent-Lock FloRest on the expiratory limb. With closure of the FloRest, the PEEP increases.

### Vent-Lock 3DP Flow Restrictor (FloRest) in Swine

Two domestic swine were anesthetized and ventilated using the Vent-Lock system with constant volume delivery. Swine were successfully ventilated for approximately four hours using a single ventilator. Initial calculated dynamic lung compliances were 50.3 and 48.1 ml/cm H_2_O for the standard and variable swine, respectively. Throughout the experiment, minimum and maximum dynamic compliance ranged from 50.3 to 244.5 and 37.9 to 87.1 ml/cm H_2_O for the standard and variable swine, respectively, reflecting the differences in tidal volumes those swine received during the flow restriction trial. Serial ABGs were monitored (Fig. 6) and initial shared ventilator settings were determined to be too high as both swine developed a respiratory alkalosis. At approximately two hours this was corrected and pH and paCO_2_ were allowed to normalize for one hour. Vent settings were not changed following this equilibration. Over the next hour the Vent-Lock system was adjusted from fully open to fully closed, where air was still allowed to pass even when Vent-Lock is closed to prevent unintentional hypoventilation. Respiratory characteristics including tidal volume ratios, percent of total set tidal volume delivered, inspiratory pressure and tidal volume are presented in Fig. 7. While the tidal volume delivered to the variable swine decreased marginally, a substantial increase in tidal volume was noted to the standard swine (Fig. 7D), similar to what is seen in simulation center testing with ventilator on volume control mode. Arterial blood gas measurements demonstrated hyperoxia in both swine (Fig. 7C). A hypercarbic respiratory acidosis occurred in the variable swine (Figure 6A, B) as the Vent-Lock closure reached its final turn. Necropsy performed to assess for gross lung pathology showed no significant findings of all lung lobes.

**Fig. 6.**
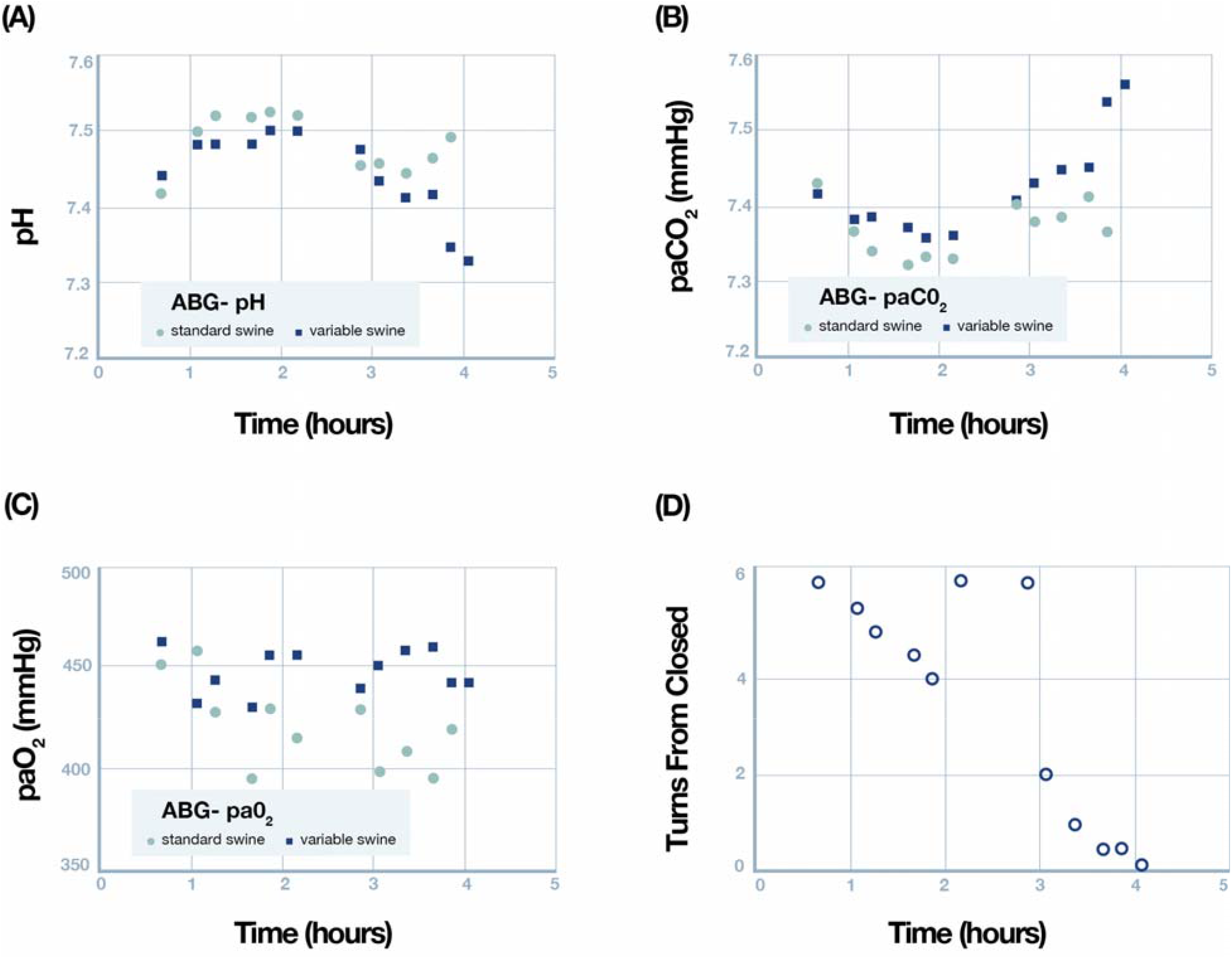
Serial Arterial Blood Gases were monitored throughout the experiment. (A) pH, (B) paCO_2_, and (C) paO_2_ are all plotted as a function of time. (D) shows the number of turns from closed (with 6 turns being fully open) as a function of the time of the experiment. Note that the Vent-Lock system was reopened at approximately 2 hours due to development of hypercarbic alkalosis.

**Fig. 7.**
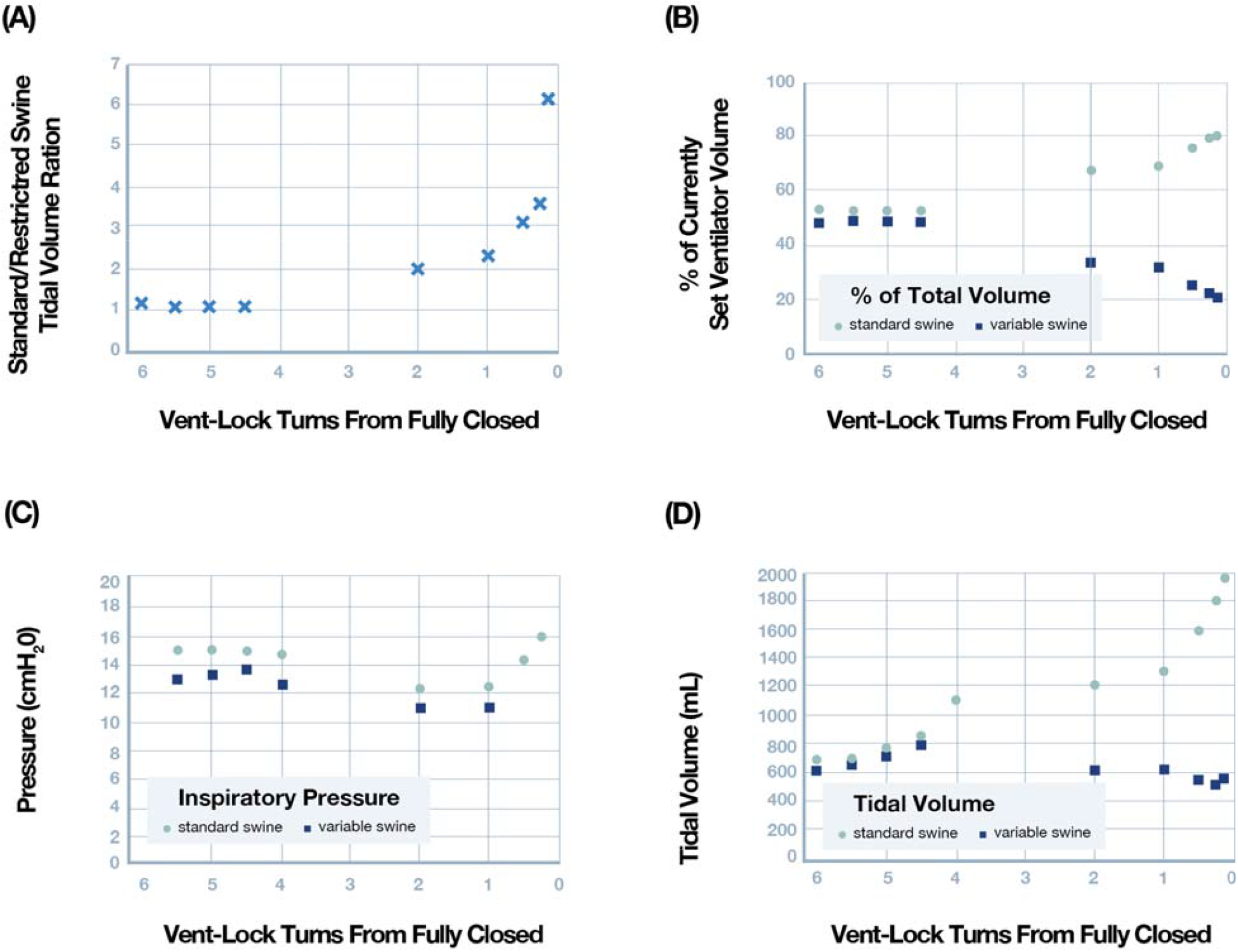
Ventilatory parameters as a function of Vent-Lock aperture with 6 turns indicating fully open and zero fully closed. The lowest turn plotted is 0.25 turns from fully closed. (A) Ratio of tidal volumes between the standard and variable swine. (B) Percent of currently set ventilator tidal volume measured in each swine. (C) Pea inspiratory pressure measured in each swine. (D) tidal volume measured in each swine.

## DISCUSSION

In the face of the COVID-19 pandemic, the importance of ventilators in the treatment of COVID-19 patients and the tenuous global supply of chain has resulted in the urgent need to increase ventilator capacity. One such solution to lack of ventilators is the ability to use one ventilator to support multiple patients. Previously reported challenges include inability to independently control flow and pressure to each patient, development of a closed system to prevent viral contamination, and continuous monitoring.

We used 3D-printing to rapidly prototype components of a ventilator splitter circuit that allows one ventilator or anesthesia gas machine to ventilate two patients. The circuit addresses aforementioned challenges and allows the clinician to control individual patient’s pressures and volumes, in a closed system with bacterial filters to reduce viral contamination. 3D printing was selected as the prototyping and production modality due to rapid iterative production for research and development and on-demand manufacturing to meet urgent needs in context of biocompatible and sterilizable 3D printing materials. 3D printed components of the splitter circuit were designed to include ventilator splitters, manometer adaptors for continuous pressure monitoring and a flow restrictor to control tidal volumes and pressures.

The novel flow restrictor valve, Vent-Lock FloRest, is able to regulate tidal volumes to one patient while maintaining flow to another patient (Fig. 3), and control PEEP when placed on the expiratory limb with the manometer adaptor (Fig. 5). Design, research, and development was driven by creating an airtight valve that allows precise control by the clinician. We demonstrated that FloRest is compatible with multiple modes of ventilation strategies and anesthesia gas machines (Fig. 4D). Vent-Lock FloRest has demonstrated volume and pressure control by different physicians, using different mechanical ventilators or anesthesia gas machines, thus demonstrating potential compatibility with different ventilators and anesthesia gas machines. While the reproducibility of the Vent-Lock FloRest must be further investigated, this has important global scalability implications as we demonstrate potential compatibility and reproducibility not limited to make or type of ventilator or anesthesia gas machines, which may be more commonly available in developing countries.

Our studies reveal fundamental differences in tidal volume patterns with flow restriction with the ventilator in volume versus pressure mode. In pressure control, Vent-Lock FloRest allowed for reduction in delivered tidal volumes to the variable patient with stable volumes delivered to the standard patient (Fig. 3A). However, in volume control, reduction in delivered tidal volume to the variable patient resulted in a concomitant increase in tidal volume delivery to the standard patient. This pattern is expected due to the continuous delivered volume maintained by the machine; therefore, the Vent-Lock FloRest allowed regulation of the ratio, the standard/variable patient tidal volume ratio (Fig. 4B). This ratio pattern of control is seen in both ventilator volume control settings in the simulation center and replicated *in vivo* swine studies using anesthesia gas machines (Fig. 5). The variation in both patients may be difficult for clinicians to manage simultaneously. However, this ratio-based control delivered by Vent-Lock FloRest can be critical for splitting legacy ventilators or anesthesia gas machines that may only have volume control settings. One of the biggest challenges of splitting patients on ventilators is that air will preferentially travel to the patient with the highest baseline lung compliance resulting in unequal ventilation between the two patients. However, if Vent-Lock FloRest is placed on the patient with the highest baseline lung compliance (variable patient), air flow can be decreased, while simultaneously increasing air flow to the standard patient, until tidal volumes are equilibrated between the two. Consequently, while we demonstrate that flow restriction provides tidal volume control on ventilator pressure or volume control settings through different mechanisms. We emphasize that especially during emergency use settings, providers appreciate these differences in tidal volume control mechanisms, and select the setting most appropriate for patients and practice settings.

It is important to note that we validated ventilation multiplexing using anesthesia gas machines. Anesthesia gas machines have ventilation functions, and are widely available globally, even in developing countries. They are well suited to be repurposed for ICU ventilation in the face of ventilator shortages, and considerations for modification and usage settings have been addressed ^24^. Our study provides follow up that in emergency situations, anesthesia gas machines can potentially be multiplexed to ventilate multiple patients using the Vent-Lock 3DP circuit.

In companion with the Vent-Lock FloRest, we also produced a manometer adaptor (Fig. 5A) that can fit standardized disposable manometers commonly found in hospital settings, such as the Ambu Disposable Pressure Monitor (Ambu, Copenhagen, Denmark). When the manometer adaptor is placed in the Vent-Lock circuit, it allows for the continuous monitoring of pressures. This is particularly important as the ventilator reported values may not accurately reflect conditions of both patients and the rapidly changing states of both patients need to be monitored with subsequent adjustments to pressures. We do note that while the manometer adaptor with the manometer provides accurate readings (Fig. 5B), it is limited by the sensitivity of the manometers for clinical use. Furthermore, using the manometer and adapter allows for real time monitoring and facilitates individual control of PEEP with use of the Vent-Lock FloRest. When the FloRest is placed on the expiratory loop of the patient, restriction of airflow results in pressure increases between the patient and the FloRest, effectively functioning as PEEP (Fig. 5C), which can then be reported by the manometer and adapter. This PEEP change established by FloRest and continuous monitoring does not affect the other patient split on the ventilator. Therefore, this is a critical asset of the circuit that allows for more patient-tailored PEEP therapy which is especially important in the treatment of patients with ARDS due to COVID-19 or other lung pathologies. While most PEEP valves currently rely on a spring-loaded control system, this may be difficult to produce rapidly, especially via additive manufacturing. Our design demonstrates control of expiratory pressures through flow restriction. However, we do note challenges with FloRest in creating PEEP control, including that the PEEP was not changed until near complete occlusion of the valve, at which point additional turns resulted in rapid changes in PEEP (Fig. 5C). Consequently, we recognize that Vent-Lock FloRest requires further optimization prior to clinical usage, but exists as a proof-of-concept that PEEP control may be possible through a spring-less system.

Some limitations to our study include lack of human testing. While the swine in this study had similar lung compliances which allowed us to show that the device was able to restrict flow and that there is a finite tidal volume that is necessary and it is unknown how well this reflects human physiology. In addition to urgent need, challenges in continuous monitoring must be addressed prior to human studies. Future directions include developing a more rigorous continuous monitoring of flow rates and delivered tidal volumes to patients to facilitate adjustments of flow per FloRest. This is critical due to the dynamic lung physiologies of patients with ARDS and preventing barotrauma or under or over ventilation. Therefore, we recommend setting a target lung volume per patient, and monitoring via spirometry or airflow transducers, such as the ones used in our swine studies (SS11LB airflow transducer (Biopac; Goleta, CA)). Patient lung volumes and their oxygenation statuses should be spot checked with the spirometer or transducers and arterial blood gases. Lastly, we emphasize that ventilator multiplexing is only to be used in emergency situations after all alternatives have been exhausted. Despite our findings of improved ventilator multiplexing functions with Vent-Lock and Vent-Lock FloRest, additional studies are required to validate the safety and clinical considerations prior to translation to human subjects. However, as future pandemics and disasters may exhaust standard-of-care for patient ventilation, Vent-Lock exists as a solution if “the other option is death” ^25^.

During the COVID-19 pandemic, open sourcing has been used to expedite the creation of vital personal protective equipment (PPE) by sharing files for the creation of masks, face shields and ventilator adjuncts. Open sourcing and 3D printing have been proven to be helpful in the developing world by providing low cost, easy to use medical products, low cost construction of homes, water treatment devices and prosthetic limbs ^26^. Thus, utilization of 3D printing to produce Vent-Lock circuit and Vent-Lock FloRest allows for the rapid, on-demand, on-site production to meet immediate needs. However, we recognize that our recommendation to use specific medical grade materials that are easily sterilized can limit production in the developing world. Further tests should be performed in varying material types to ensure accuracy in the printing process and translation into actual use. While it is promising that our materials have remained stable in humidified 40 °C for over 48 hours (fig. S3), we note that the material appears more brittle after multiple autoclave cycles, and thus further testing is required to ascertain stability across pressure gradients over weeks to months of use. Thus, we recommend single use of the devices until further investigation.

Additionally, we do recognize that the value of Vent-Lock circuit is also in its ability to be stored in preparation of emergency situations, such as disaster preparedness or in military combat zones, where ventilator shortages can be expected. In this case, we believe that while 3D printing production can meet initial interests, traditional manufacturing (such as injection molding), may be a more cost-effective and time-efficient approach to fulfil demand. While Vent-Lock circuit components are not currently optimized for traditional manufacturing, the designs can easily be modified to allow for this production modality, while still maintaining access to 3D print for instances where manufacturing infrastructure is unavailable to produce this device. Consequently, we believe that open sourced 3D printing methodology of Vent-Lock production is appropriate for scalability in the face of urgent demand, such as the COVID-19 pandemic, but can also be transitioned into traditional manufacturing.

In this study, we developed Vent-Lock, a ventilator or anesthesia gas machine splitter system with a flow restrictor (FloRest) that can modify flow rates per patient for patient tailored therapies. We provide proof-of-concept that two swine can be safely split using one anesthesia gas machine. While additional work is critical for the safe use of ventilator multiplexing, our data supports the feasible use of ventilator splitting for emergency situations, such as in face of COVID-19.

## MATERIALS AND METHODS

### 3D printing procedure

3D printing of the Vent-Lock splitters, flow regulators, and manometer adaptors were produced via stereolithography (Form 2, Form 3, or Form 3B, Formlabs) at 50 um layer resolutions, using surgical guide resin (Surgical Guide, Formlabs). Print files were generated by CAD drawings (SolidWorks, Dassault Systèms) and converted into G-code using the printer’s accompanying software package (PreForm, Formlabs). Support structures were minimized through design and generated using PreForm where needed. Components were oriented in such a way that crucial surfaces such as threads or O-ring ledges were not impacted by support structures. Prints were post-processed by washes (2 cycle with 15 min per cycle) in >99.5% isopropyl alcohol (CAS Number: 67-63-0, Sigma Aldrich), followed by air-drying at 22 °C for 30 minutes, and post-cured for 30 minutes with heat 60 °C for the Form 2 printer and 70 °C for the Form 3B printer at 405 nm of light (Form Cure, Formlabs). O-rings (E1000-212/AS568-212, O-Rings EPDM, FDA EPDM, Marco Rubber &Plastics, Seabrook, New Hampshire, USA) were added for improved sealing. Production via fused deposition modeling (FDM) (e3d, BigBox3D Ltd, Oxfordshire, UK; Little Monster, Tevo 3D Electronic Technology Co. Ltd, Zhanjiang, China) used PETG Filament (PETG 3D Printer Filament, FilaMatrix, Virginia, USA). Print settings were a 0.2mm layer height with 30% infill, nozzle temperature of 250 °C, and bed temperature of 70 °C; supports were generated from the build platform, with no interior supports.

### Sterilization Testing

3D-printed parts produced from surgical guide resin were sterilized by dry vacuum autoclave (Sr 24C Adv-Plus™, Consolidated Sterilizer Systems, Boston, Massachusetts, USA), 3 cycles at 120.0 oC, 20 minutes sterilization time and 20 minutes dry time. Then, they were soaked in >99.5% isopropyl alcohol (CAS Number: 67-63-0, Sigma Aldrich) for 30 minutes, air-dried at 22 oC for 30 minutes, and placed in an oven at 40 °C in humidified air for 48 hours (VO1824HPC, Lindberg/Blue M Vacuum Oven 127.4L, Thermo Scientific, Waltham, MA, USA). Particle count analyses were conducted using a particle counter (SOLAIR 3100, Lighthouse Worldwide Solutions), detecting sizes 0.3 to 10 microns, for 1-minute cycles, and performed for parts pre-autoclave, post-autoclave and post IPA wash, and humidified warm air exposure at 40 °C.

#### Vent-Lock 1+n(1) circuit and components

Vent-Lock circuits were assembled as depicted in Fig. 1. Vent-Lock 3DP splitters, flow regulator, and manometer adaptors were used. Commercial components include manometer (Ambu Disposable Pressure Manometer, Ambu, Copenhagen, Denmark), one-way valves (22F x 22M, REF 50245, Mallinckrodt Pharmaceuticals), disposable bacteria filters (BSF104, Vincent Medical), and ventilator tubing (SKU: 999027588, Hudson Rci).

#### Simulation Center Testing

Vent-Lock 1+n(1) circuits were tested at the Johns Hopkins Medicine Simulation Center (JHMSC). The ventilator (Puritan Bennett 840 Ventilator System, Avante Health Solutions) was using pressure control mode of ventilation (Volume Ventilation Plus™, Avante Health Solutions) with additional settings detailed in fig. S4. Vent-Lock 1+1 circuit was tested using test lungs simulating healthy lungs with variable compliances (Standard patient: R_p_ = 2 cmH_2_O/L/s, RespiTrainer Advance, QuickLung, IngMar Medical; Variable patient: R_p_= 50 cmH_2_O/L/s, ASL 5000, IngMar Medical). Intrapulmonary data for both patients were collected; data included peak inspiratory pressures, tidal volumes, and peak end expiratory pressures. Five total values of tidal volume per data set were collected and averaged. Corresponding ventilator reported data was also recorded, including total expiratory volumes, peak inspiratory pressures, mean inspiratory pressures, and peak end expiratory pressures. Flow restrictors (#P20034 PVC SCH 40 ½-in FNPT Ball Valve; G300 Lead Free Brass Gate Valve; #P60SCPVC12 Stop and Waste Valve, American Valve, Greensboro, North Carolina, USA; Vent-Lock 3DP FloRest) were used to restrict the variable patient’s inspiratory flow rate per the 1+n(1) circuit (Fig. 1). Valve handles were turned at smallest increments permissible to close the valve and documented as % closure. Corresponding intrapulmonary data and ventilator reported values were collected per handle closure and standardized to values (volumes and pressures) of a fully open valve (reported as proportion of maximum, %).

In vitro studies were also conducted at the Washington University Simulation Center using two Datex-Ohmeda Aestiva anesthesia machines. One machine was set to deliver pressure control ventilation in a manner similar to that performed at Johns Hopkins. This machine was connected in parallel to a 2L anesthesia bag reservoir and a second Datex-Ohmeda Aestiva machine that was set to spontaneous ventilation. The second machine served as a flow and volume sensor for the Vent-Lock 1+n(1) circuit.

### *In vivo* swine studies

Experiments were performed in accordance with the Guide for the Care and Use of Laboratory Animals and were approved by the Institutional Animal Care and Use Committee of Washington University School of Medicine (St. Louis, MO). Domestic swine (*Sus scrofa domesticus*) were purchased from Oak Hill Genetics (Ewing, IL). The swine were females, 72 kg each, 5 months old, and were Landrace-cross swine. Swine were sedated with a telazol, ketamine, xylazine cocktail and intubated with a 7.0 endotracheal tube. Anesthesia was maintained with isoflurane. Femoral venous and arterial catheterization was performed. Standard ASA monitoring was maintained throughout the experiment. Swine were ventilated using a single ventilator (Drager Narkomed 2A) with two circuits in parallel in an 1+n(1) configuration with cross-ventilation restricted by using one-way check valves. Ventilation was maintained with volume control. One swine was not flow-regulated and thus considered the standard patient, while the other had a Vent-Lock 3DP 4.0 connected in the inspiratory limb and thus considered the variable patient. Flow was measured at each expiatory limb with a SS11LB airflow transducer (Biopac; Goleta, CA). Flow data were collected at 2kHz using an MP36 data acquisition unit and BSL 4.1.3 software (Biopac; Goleta, CA). The spirometry data was then smoothed with a 0.25 sec wide moving median filter after removal of instrument noise below 0.08 L/sec (determined by histogram inspection). The smoothed data was then numerically integrated to estimate respiratory tidal volume, and a first order numeric derivative was used to calculate the instantaneous respiratory rate. The noise floor for the integrated volume was determined by histogram inspection resulting in a threshold of 90 mL. The anesthesia record and the spirometry results were then aligned using common timestamps. All breaths spontaneously initiated by the swine (identified by respiratory rates more than 30% away from the ventilator set point) were removed from analysis. The mean and standard deviation for each anesthesia record entry were calculated for respiratory rate, tidal volume, minute ventilation, and lung compliance. All of the described analysis was performed using a custom MATLAB script (MATLAB 2019b, The MathWorks, Inc., Natick, MA)]. Arterial and venous blood gas data were collected 15 minutes following any changes to the Vent-Lock 3DP device. Following the procedure, swine were euthanized with an overdose (∼150mg/kg) of supersaturated potassium chloride IV while under anesthesia. Necropsy was performed to assess for any gross lung pathology.

## Data Availability

Data is available upon request

## ACKNOWLEDGEMENTS

We would like to thank the Johns Hopkins Medicine Simulation Center Jordan Duval-Arnould, Julie Perretta, and Joe Dwyer for use of the simulation center; Dr. Thao (Vicky) Nguyen, Dr. Joe Katz for their advice; Dr. Adam Sapirstein for their medical expertise; Dr. Sarah Clever for her support; the Johns Hopkins Applied Physics Lab for guidance and advice; Dr. Gerald Brandacher and Yi-nan Guo for use of their autoclave; Gregory Bova for loan of his particle counter; and Dr. Arun Agrawal and Azra Horowitz for their advice on ventilator splitting. The authors acknowledge financial support from the Johns Hopkins University President’s Response to COVID-19 Fund, the Start-Up Fund from the Whiting School of Engineering at Johns Hopkins University, support from the Department of Civil and Systems Engineering and Johns Hopkins Center for Additive Manufacturing and Architected Materials, and the National Science Foundation (DMREF-1628974). JU acknowledges financial support from the U.S. Army Research Office (ARO) sponsored NDSEG Fellowship program and AT acknowledges support by a NASA Space Technology Research Fellowship.

## AUTHOR CONTRIBUTIONS

HX, JKG, SHK, and JC conceived the research. HX, CS, JU, RHT, AT, MV, JF, MS, BW, JKG, SHK, and JC contributed to engineering designs and production. HX, CS, JU, RT, AT, MV, JF, MS, PY, BSK, BW, JKG, SHK, and JC contributed to experimental design and testing in the JHMSC. BH and JF contributed to communications and graphic design. BB, MM, MT, MF, BK, and JMS contributed to simulation center testing and swine studies at Washington University in St. Louis. JKG, SHK, and JC supervised the research. All authors contributed to the drafting and editing of this manuscript.

## SUPPLEMENTARY MATERIALS

Fig. S1. *De novo* ventilator circuit components produced via 3D printing

Fig. S2. Air-tightness tests of the Vent-Lock FloRest.

Fig. S3. Design files for Vent-Lock splitters, needle valve, and manometer adaptor.

Fig. S4. Tests of tidal volume control with and without O-rings.

Fig. S5. Comparisons of Vent-Lock FloRest performances depending on materials.

Table S1. The biodurability and sterilization conditions.

Table S2. Ventilator settings on 840 Ventilator System, Nellcor Puritan Bennett.

Mov S1. Leaky bubble test demonstrates Vent-Lock FloRest is airtight

Data S1. STL print files of Vent-Lock splitter, FloRest, and manometer adaptor

Data S2. MATLAB code used to analyze swine data

